# Social Determinants of Health and Long COVID in U.S. Children: A Cross-Sectional Study, 2022--2023

**DOI:** 10.64898/2026.07.29.26359222

**Authors:** Douglas Slaughter, Krysten Rose-McCully, Hope King, Caroline Pratt, Alexandra F. Rollins, Sharon Saydah, Nicole D. Ford

## Abstract

**Objective:** We characterized social determinants of health (SDOH) in U.S. children who ever and never had Long COVID.

**Methods:** We used cross-sectional data from the 2022-2023 National Health Interview Survey (N=14,993 children 0-17 years). Parents reported child- and household-level information. Long COVID was defined as ever experiencing symptoms lasting ≥3 months that were not present prior to having COVID-19. We produced weighted prevalence estimates by Long COVID status for 3 SDOH domains (Social and Community Context, Healthcare Access and Quality, and Economic Stability) and used Rao-Scott chi-squared tests to examine differences.

**Results:** In Social and Community Context, children who ever had Long COVID more often resided in single parent households (33.2% vs. 20.3%; p<0.0001), with someone with severe depression or mental illness (18.5% vs. 8.3%; p<0.0001) or substance abuse (16.7% vs. 8.1%; p<0.0001) or had a lifetime of being disparaged by adults in the home (9.2% vs. 3.9%; p=0.0009). In Healthcare Access and Quality, children who ever had Long COVID more often had public insurance (e.g., Medicaid) (51.3% vs. 41.6%; p=0.02), higher healthcare use, and difficulty paying medical bills (23.2% vs. 12.2%; p<0.0001). In Economic Stability, children who ever had Long COVID had lower parental education, lower food security, and higher participation in social safety net programs (p<0.01 for all comparisons).

**Conclusion:** Children who ever had Long COVID more frequently experienced adverse SDOH compared to their peers who never had Long COVID. These findings may help identify children who may benefit from additional resources related to their Long COVID care.

**Article Summary:** Children who had Long COVID more frequently reported adverse Social Determinants of Health (SDOH) compared to their peers who never had Long COVID.

**What’s Known on This Subject:** The relationship of SDOH and Long COVID in children has not been well characterized; however, research indicates potential disparities in Long COVID symptoms and severity based on age, sex, race, ethnicity, and socioeconomic characteristics.

**What This Study Adds:** In a nationally representative sample, we found that U.S. children who had Long COVID more frequently reported adverse SDOH. These results may help identify higher-risk children who may benefit from additional resources related to their Long COVID care.

## Background

Long COVID is a chronic condition that includes a wide range of prolonged symptoms lasting at least 3 months following SARS-CoV-2 infection.^1^ During 2023, 0.4% of U.S. children had Long COVID and 1.4% had ever had Long COVID. Among children who experienced Long COVID during 2023, 80% reported activity limitations.^2,3^ Risk factors for Long COVID in pediatric populations include older age (i.e., adolescence compared to younger aged children), female sex, more severe acute COVID-19 illness, obesity, and preexisting chronic conditions.^4,5^

Social determinants of health (SDOH) are conditions in the environments where people are born, live, learn, work, play, worship, and age that affect a wide range of health, function, and quality of life outcomes.^6^ Examples of SDOH include frequency of healthcare access, stressful life events, healthcare affordability, annual household income, educational attainment, access to clean water, and safe housing.^6,7^ SDOH can affect the likelihood that someone develops or is diagnosed with a medical condition and one’s ability to seek and manage care for medical conditions such as Long COVID. Studies examining the association between SDOH and Long COVID in U.S. adults reported that adults with Long COVID were roughly 1.5 times more likely to experience significant difficulty with household expenses and 1.9 times more likely to face eviction or foreclosure compared to adults without Long COVID.^8–10^ Competing priorities such as consistent shelter and affordable food for the household can influence whether a person is able to adequately manage their medical care. This may be particularly true for children and adolescents who are reliant on caregivers to navigate health care and educational systems on their behalf.

The role of SDOH and Long COVID in children has not been well characterized.^11^ Limited research has indicated potential differences in pediatric Long COVID symptoms and symptom severity by age, sex, race, ethnicity, and other socioeconomic characteristics. For example, in an electronic health record-based cohort study, children aged younger than 5 years and Hispanic children both had higher odds of having respiratory Long COVID symptoms while non-Hispanic white children and those 10 years of age and older had higher odds of neurologic Long COVID symptoms.^12^ However, findings are sparse; SDOH in children with Long COVID was highlighted as a significant gap in the current literature.^11^

In this analysis, we descriptively characterized selected household- and child-level SDOH indicators among U.S. children with and without a caregiver-reported history of Long COVID. Although cross-sectional data prevents us from determining a causal relationship between SDOH and Long COVID, better understanding these associations could help identify children who may benefit from monitoring their acute COVID-19 episode or additional resources to address Long COVID.

## Methods

### Study Population

We combined and analyzed data from the 2022 and 2023 National Health Interview Survey (NHIS). The NHIS is a nationally representative cross-sectional survey that collects health information from the non-institutionalized civilian U.S. population.^13^ In multi-staged sampling, NHIS first identifies households. Among sampled households, one child aged 17 years or younger is randomly selected to participate. The child’s parent or knowledgeable adult (henceforth referred to as caregiver) responds to interviewer-administered questionnaires.^13,14^ Details about survey methodology are publicly available along with the survey data. ^13,14^

### COVID-19 and Long COVID

Children were classified as having a history of COVID-19 if their survey responses indicated they took a test confirming the presence of COVID-19 or if they were told by a medical professional they had (or likely had) COVID-19. For children with prior COVID-19 illness, caregivers reported whether children had any symptoms lasting 3 months or longer that were not present prior to having COVID-19 illness. Those responding affirmatively to the question about long term symptoms were classified as ever having Long COVID. Children who never had COVID-19 and those with prior COVID-19 who did not have symptoms that lasted 3 months or longer were classified as never having Long COVID. Because national surveys underestimate prior COVID-19 compared to seroprevalence surveys, we did not analyze children without prior COVID-19 separately.^15^ This analysis describes SDOH among children who ever had Long COVID; the sample of children with current Long COVID was too small to present reportable estimates.

### Social Determinants of Health

The U.S. Department of Health and Human Services (HHS) organizes SDOH into 5 domains.^6^ The Social and Community Context domain addresses relationships and interactions with family, friends, co-workers, and community members, social support, and Adverse Childhood Experiences (ACEs) - traumatic events that occur in childhood affecting an individual over the life course.^6^ The Healthcare Access and Quality domain addresses impediments to and factors that support the timely receipt of quality healthcare (e.g., adequate health insurance coverage, physical proximity to health centers, and having a primary care provider).^6^ Economic Stability encompasses factors such as poverty, employment, and the ability to afford healthy foods, healthcare, and housing.^6^ Education Access and Quality addresses education, including education quality.^6^ Lastly, Neighborhood and Built Environment includes indicators capturing the impact of “place,” such as availability of healthy food and safe water, neighborhood violence, clean air, sidewalks, and green spaces.^6^

Two authors independently reviewed relevant survey questions from the NHIS child survey and grouped them according to the HHS SDOH framework. Disagreements were managed through discussion and co-author consensus. Based on the survey questions in NHIS, we identified available variables for 3 of the 5 domains (Social and Community Context, Healthcare Access and Quality, and Economic Stability) and focused our analysis on these indicators. We also included other child-level sociodemographic characteristics such as age, sex, race, and ethnicity. A complete table of all indicators identified for this analysis grouped by domain is presented in Table 1.

**Table 1.**
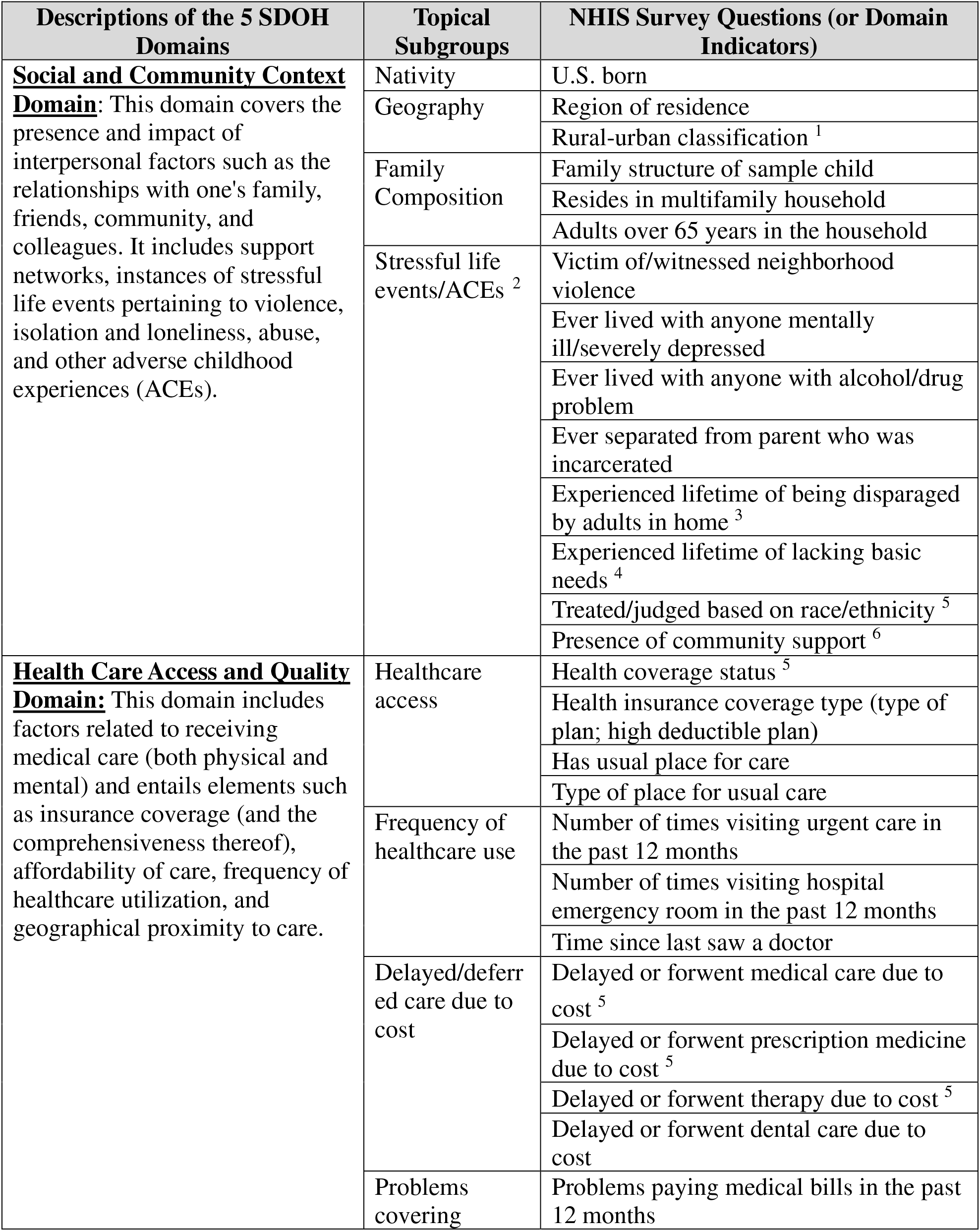
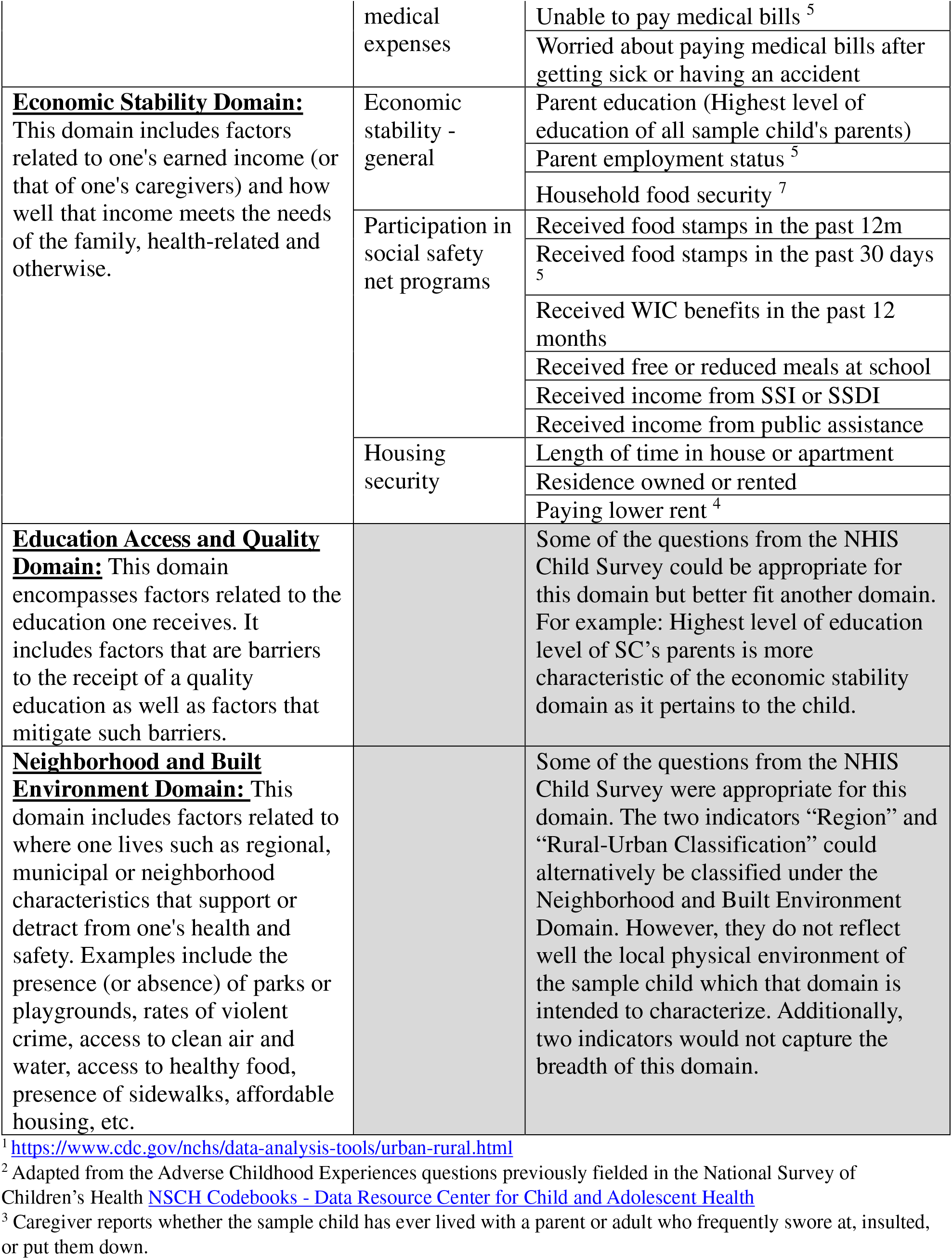
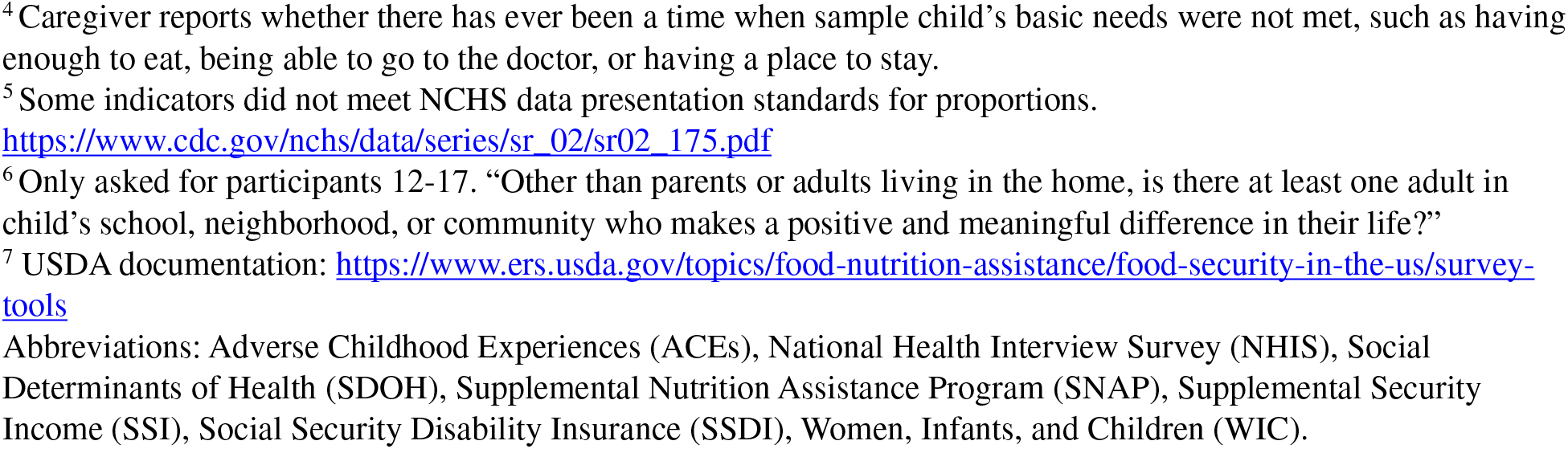
Organization of National Health Interview Survey (NHIS) child survey questions into the 5 domains of the Social Determinants of Health (SDOH) framework.

### Statistical Methods

We produced weighted prevalence estimates (95% Clopper-Pearson Confidence Intervals [CI]) for all domain indicators for children who ever and never had Long COVID and used Rao-Scott chi-squared tests to examine differences by Long COVID status.

Analyses were conducted using SAS v.9.4 and SAS-callable SUDAAN. We used NHIS-supplied sampling weights, domain analysis, and survey procedures to account for complex sampling in the NHIS. Two-sided P-values <0.05 were considered statistically significant. Estimates not meeting NCHS standards for reporting proportions were not presented.^16^

This work was reviewed by CDC, deemed not research, and was conducted consistent with applicable federal law and CDC policy (45 C.F.R. part 46.102(l)(2), 21 C.F.R. part 56; 42 U.S.C. Sect. 241(d); 5 U.S.C. Sect. 552a; 44 U.S.C. Sect. 3501 et seq.).

## Results

### Study Sample

During 2022-2023, 15,156 children 0-17 years were sampled (N=7,464 in 2022 and N=7,692 in 2023). We excluded participants who were missing information about COVID-19 history (N=143) and those for whom Long COVID status was unknown (N=20). Our final unweighted analytic sample was 14,993 children (N=71,622,000 weighted).

Based on the weighted sample, an estimated 1.3% (unweighted n=212) children reported ever having Long COVID. Female children and adolescents aged 12-17 years had disproportionately higher reported Long COVID (58.6% and 55.5%, respectively, compared with 41.4% of male children and 44.5% of children aged 0-11 years of age) (Data not shown). Children with Hispanic ethnicity were also disproportionately affected, with 37.5% having ever had Long COVID compared with 25.9% of those who never had Long COVID.

### Social and Community Context

Children who ever and never had Long COVID reported differed on Social and Community context domain indicators. Compared to children who never had Long COVID reported, children who ever had Long COVID reported more frequently resided in a single parent household (33.2% compared with 20.3%, p<0.0001), lived with someone with severe depression or mental illness (18.5% compared with 8.3%, p<0.0001), lived with someone with alcohol or drug problems (16.7% compared with 8.1%, p=0.0001), or had a lifetime of being disparaged by adults in the home (9.2% compared to 3.9%, p=0.0009) (Table 2).

**Table 2.** Social and Community Context Domain indicators among children who ever and never had Long COVID, United States, 2022-2023. ^1^

| Characteristic | Percent of Children who ever had Long COVID |  | Percent of Children who never had Long COVID |  | P-value <sup>2</sup> |
| --- | --- | --- | --- | --- | --- |
|  | N | (95% CI) | N | (95% CI) |  |
| **Total** | 212 | 1.3 (1.2, 1.6) | 14,781 | 98.7 (98.4, 98.8) |  |
| **Nativity** |  |  |  |  |  |
| U.S. born | 204 | 96.8 (93.0, 98.8) | 13,864 | 95.3 (94.8, 95.8) | 0.37 |
| **Geography** |  |  |  |  |  |
| U.S. Census region of residence <sup>3</sup> |  |  |  |  | 0.83 |
| Northeast | 25 | 14.4 (9.1, 21.1) | 2,175 | 15.9 (14.6, 17.2) |  |
| Midwest | 47 | 20.7 (15.2, 27.1) | 3,050 | 20.9 (19.5, 22.4) |  |
| South | 78 | 38.1 (30.7, 45.9) | 5,515 | 39.3 (37.3, 41.3) |  |
| West | 62 | 26.8 (20.6, 33.8) | 4,041 | 23.9 (22.1, 25.8) |  |
| Urban-Rural Classification <sup>4</sup> |  |  |  |  | 0.73 |
| Large Central Metropolitan | 60 | 28.3 (21.3, 36.1) | 4,542 | 29.6 (27.3, 32.0) |  |
| Large Fringe Metropolitan | 50 | 23.9 (17.1, 31.8) | 3,800 | 26.6 (24.1, 29.1) |  |
| Medium and Small Metropolitan | 71 | 32.5 (25.4, 40.2) | 4,478 | 30.6 (27.6, 33.8) |  |
| Non-metropolitan | 31 | 15.3 (10.1, 21.9) | 1,961 | 13.2 (11.9, 14.6) |  |
| **Family composition** |  |  |  |  |  |
| Family structure |  |  |  |  | <0.0001 |
| Single parent | 76 | 33.2 (26.3, 40.7) | 3,095 | 20.3 (19.4, 21.3) |  |
| Two parents living in same household | 108 | 53.9 (46.0, 61.8) | 9,962 | 68.4 (67.3, 69.5) |  |
| Other or unknown | 28 | 12.9 (8.3, 18.7) | 1,699 | 11.3 (10.7, 11.9) |  |
| Multifamily household | 2 | 0.8 (0.1, 3.2) | 142 | 1.0 (0.8, 1.2) | 0.85 |
| Adults over 65 years in the household | 16 | 8.2 (4.3, 14.0) | 1,103 | 6.5 (6.1, 7.0) | 0.41 |
| **Stressful life events/ACEs** |  |  |  |  |  |
| Victim of/witnessed violence | 17 | 5.9 (3.1, 10.0) | 781 | 5.5 (5.1, 6.1) | 0.80 |
| Ever lived with anyone mentally ill/severely depressed | 40 | 18.5 (13.2, 24.9) | 1,174 | 8.3 (7.7, 8.9) | <0.0001 |
| Ever lived with anyone with alcohol/drug problem | 35 | 16.7 (11.0, 23.7) | 1,181 | 8.1 (7.5, 8.7) | 0.0001 |
| Ever separated from parent who was incarcerated | 22 | 9.4 (5.4, 14.9) | 835 | 6.1 (5.6, 6.7) | 0.08 |
| Lifetime of being disparaged by adults in home <sup>5</sup> | 21 | 9.2 (5.2, 14.9) | 562 | 3.9 (3.5, 4.4) | 0.0009 |
| Lifetime of lacking basic needs <sup>6</sup> | 6 | 2.3 (0.8, 5.4) | 473 | 3.3 (3.0, 3.7) | 0.41 |
| Presence of community support <sup>7</sup> | 104 | 84.9 (74.9, 92.1) | 4,726 | 88.0 (86.8, 89.2) | 0.41 |

<sup>1</sup> Ns are unweighted. Estimates are % (95% CI) and are weighted and account for complex sampling. “Treated or judged based on race/ethnicity” is not presented because estimates did not meet NCHS data presentation standards for proportions
(https://www.cdc.gov/nchs/data/series/sr_02/sr02_175.pdf)
<sup>2</sup> P-values are for Rao-Scott Chi Square test of independence for each domain indicator by Long COVID status.
<sup>3</sup> https://www.cdc.gov/nchs/hus/sources-definitions/geographic-region.htm
<sup>4</sup> https://www.cdc.gov/nchs/data-analysis-tools/urban-rural.html
<sup>5</sup> Caregiver reports whether the sample child has ever lived with a parent or adult who frequently swore at, insulted, or put them down.
<sup>6</sup> Caregiver reports whether there has ever been a time when sample child’s basic needs were not met, such as having enough to eat, being able to go to the doctor, or having a place to stay.
<sup>7</sup> Only asked for participants aged 12-17. “Other than parents or adults living in the home, is there at least one adult in child’s school, neighborhood, or community who makes a positive and meaningful difference in their life?”
Abbreviations: Adverse Childhood Experiences (ACEs), Confidence Interval (CI)

### Health Care Access and Quality

Having *any* health insurance coverage (i.e., insured vs. uninsured) did not vary by Long COVID status; however, type of coverage (private, public, uninsured) did (Table 3). Among children who ever had Long COVID, 51.3% had Medicaid coverage or another publicly-funded insurance compared with 41.6% of those who never had Long COVID (p=0.02).

**Table 3.**
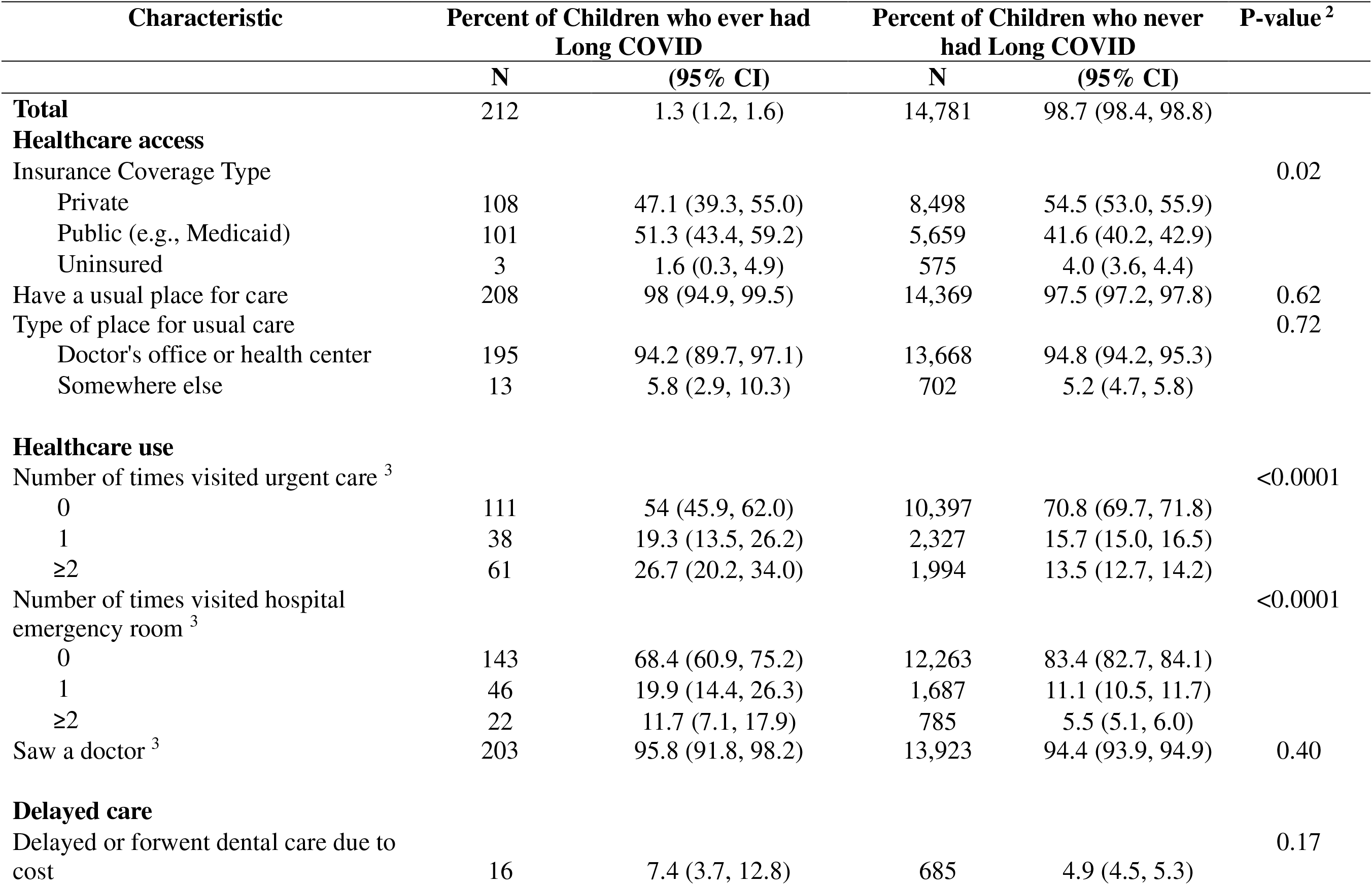
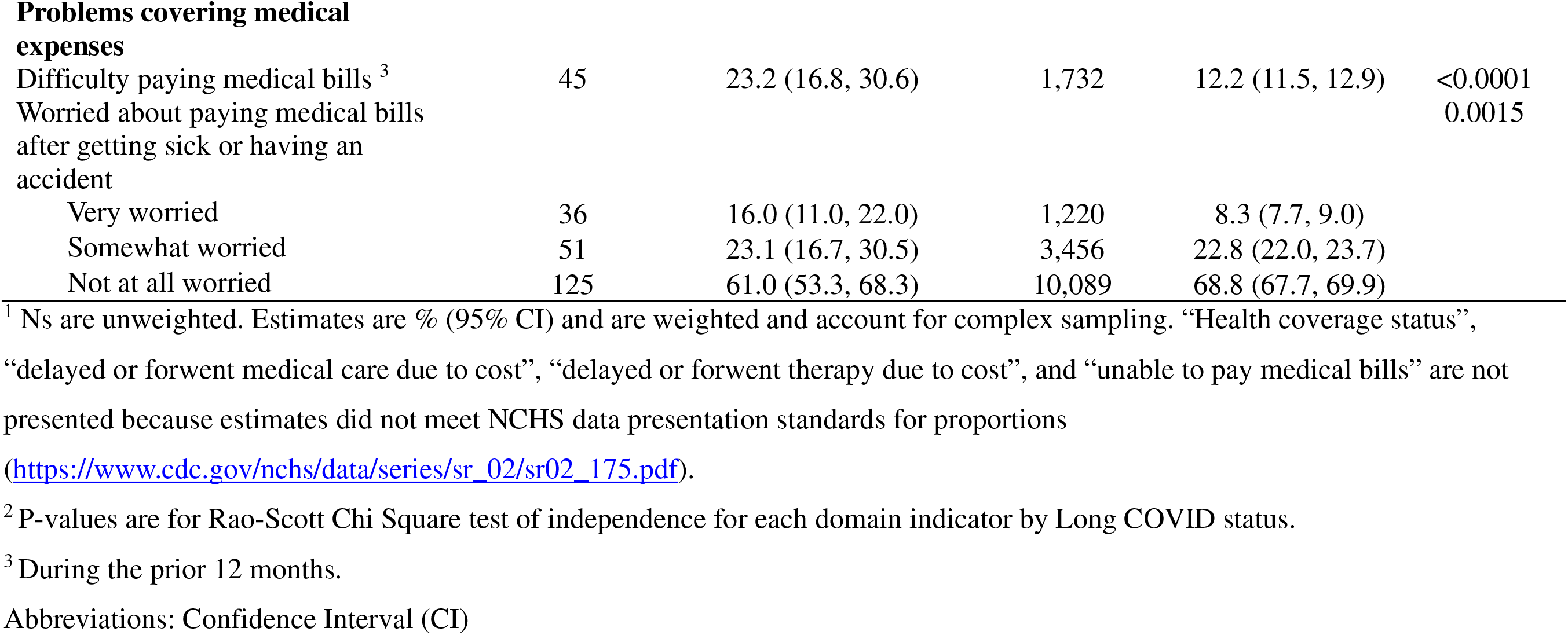
Health Care Access and Quality Domain indicators among children who ever and never had Long COVID, United States, 2022-2023. ^1^

| Characteristic | Percent of Children who ever had Long COVID |  | Percent of Children who never had Long COVID |  | P-value <sup>2</sup> |
| --- | --- | --- | --- | --- | --- |
|  | N | (95% CI) | N | (95% CI) |  |
| **Total** | 212 | 1.3 (1.2, 1.6) | 14,781 | 98.7 (98.4, 98.8) |  |
| **Healthcare access** |  |  |  |  |  |
| Insurance Coverage Type |  |  |  |  | 0.02 |
| Private | 108 | 47.1 (39.3, 55.0) | 8,498 | 54.5 (53.0, 55.9) |  |
| Public (e.g., Medicaid) | 101 | 51.3 (43.4, 59.2) | 5,659 | 41.6 (40.2, 42.9) |  |
| Uninsured | 3 | 1.6 (0.3, 4.9) | 575 | 4.0 (3.6, 4.4) |  |
| Have a usual place for care | 208 | 98 (94.9, 99.5) | 14,369 | 97.5 (97.2, 97.8) | 0.62 |
| Type of place for usual care |  |  |  |  | 0.72 |
| Doctor's office or health center | 195 | 94.2 (89.7, 97.1) | 13,668 | 94.8 (94.2, 95.3) |  |
| Somewhere else | 13 | 5.8 (2.9, 10.3) | 702 | 5.2 (4.7, 5.8) |  |
| **Healthcare use** |  |  |  |  |  |
| Number of times visited urgent care <sup>3</sup> |  |  |  |  | <0.0001 |
| 0 | 111 | 54 (45.9, 62.0) | 10,397 | 70.8 (69.7, 71.8) |  |
| 1 | 38 | 19.3 (13.5, 26.2) | 2,327 | 15.7 (15.0, 16.5) |  |
| ≥2 | 61 | 26.7 (20.2, 34.0) | 1,994 | 13.5 (12.7, 14.2) |  |
| Number of times visited hospital emergency room <sup>3</sup> |  |  |  |  | <0.0001 |
| 0 | 143 | 68.4 (60.9, 75.2) | 12,263 | 83.4 (82.7, 84.1) |  |
| 1 | 46 | 19.9 (14.4, 26.3) | 1,687 | 11.1 (10.5, 11.7) |  |
| ≥2 | 22 | 11.7 (7.1, 17.9) | 785 | 5.5 (5.1, 6.0) |  |
| Saw a doctor <sup>3</sup> | 203 | 95.8 (91.8, 98.2) | 13,923 | 94.4 (93.9, 94.9) | 0.40 |
| **Delayed care** |  |  |  |  |  |
| Delayed or forwent dental care due to cost | 16 | 7.4 (3.7, 12.8) | 685 | 4.9 (4.5, 5.3) | 0.17 |
| **Problems covering medical expenses** |  |  |  |  |  |
| Difficulty paying medical bills <sup>3</sup> | 45 | 23.2 (16.8, 30.6) | 1,732 | 12.2 (11.5, 12.9) | <0.0001 |
| Worried about paying medical bills after getting sick or having an accident |  |  |  |  | 0.0015 |
| Very worried | 36 | 16.0 (11.0, 22.0) | 1,220 | 8.3 (7.7, 9.0) |  |
| Somewhat worried | 51 | 23.1 (16.7, 30.5) | 3,456 | 22.8 (22.0, 23.7) |  |
| Not at all worried | 125 | 61.0 (53.3, 68.3) | 10,089 | 68.8 (67.7, 69.9) |  |

<sup>1</sup> Ns are unweighted. Estimates are % (95% CI) and are weighted and account for complex sampling. “Health coverage status”, “delayed or forwent medical care due to cost”, “delayed or forwent therapy due to cost”, and “unable to pay medical bills” are not presented because estimates did not meet NCHS data presentation standards for proportions (https://www.cdc.gov/nchs/data/series/sr_02/sr02_175.pdf).
<sup>2</sup> P-values are for Rao-Scott Chi Square test of independence for each domain indicator by Long COVID status.
<sup>3</sup> During the prior 12 months.
Abbreviations: Confidence Interval (CI)

Both groups reported having a usual place for medical care, similar types of places for usual medical care, and similar amounts of time since having last seen a doctor. However, children who ever had Long COVID reported had higher reported healthcare use. Compared with those who never had Long COVID reported, children who ever had Long COVID reported had ≥2 reported urgent care visits (26.7% compared to 13.5%) and ≥2 reported visits to the emergency department (11.7% compared to 5.5%) during the 12 months preceding the survey (p<0.0001).

Households of children who ever had Long COVID more frequently reported having trouble paying medical bills during the 12 months preceding the survey, with 23.2% reporting such difficulties compared to 12.2% of households of children who never had Long COVID (p<0.0001). Further, 16.0% of households of children who ever had Long COVID reported being very worried about paying future medical bills, which was nearly double the prevalence compared to households of children who never had Long COVID (8.3%) (p=0.0015).

### Economic Stability

Parental education differed by reported child Long COVID status; approximately one in three children who ever had Long COVID had parents who completed a bachelor’s degree (34.0%) compared with nearly half of children who never had Long COVID (48.0%) (p=0.0002) (Table 4). Compared with children who never had Long COVID, children who ever had Long COVID more often received supplementary income or public assistance (14.0% compared to 6.4%) and resided in households which had lower food security (10.1% compared to 4.2%) (p<0.001 for both comparisons).

**Table 4.**
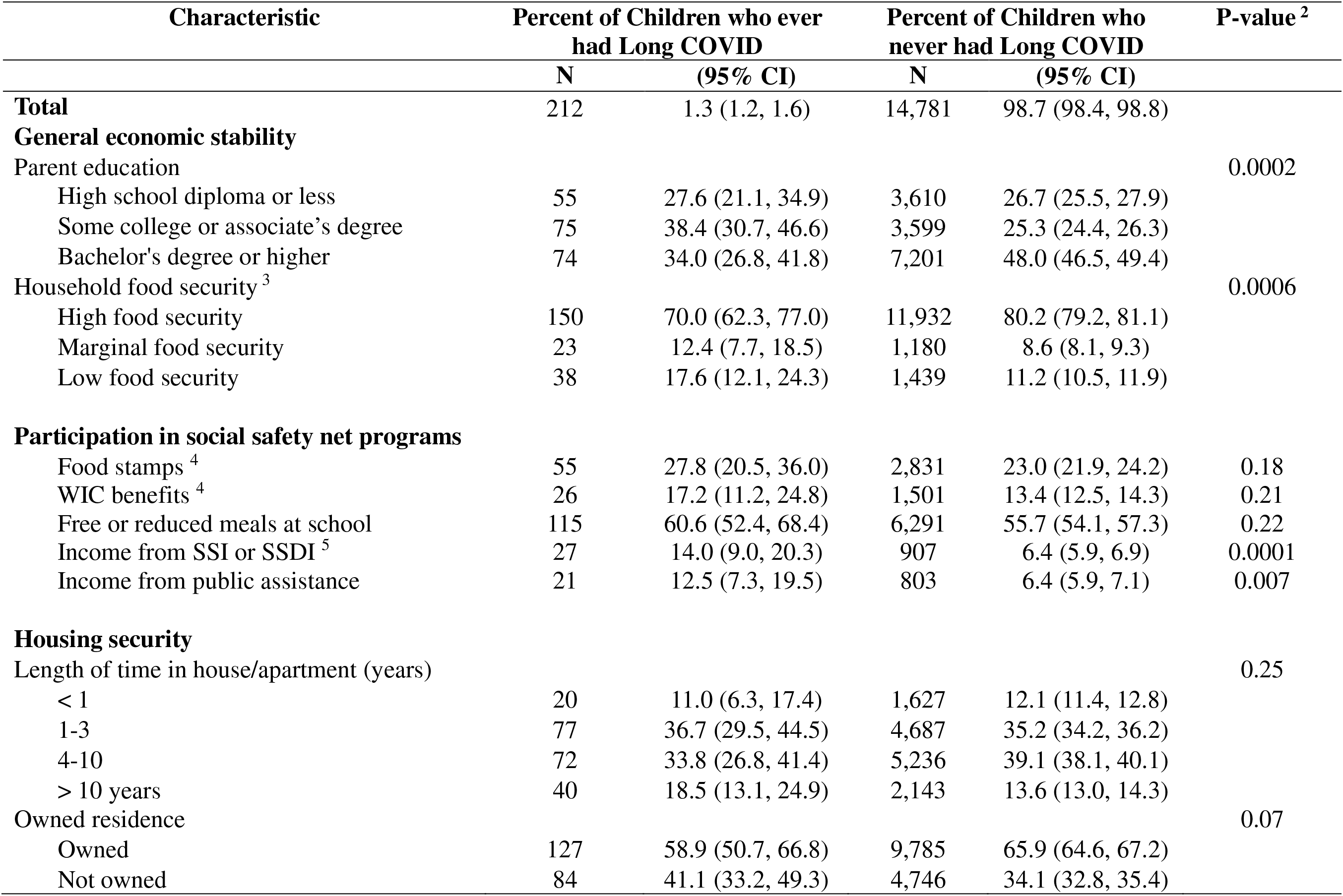
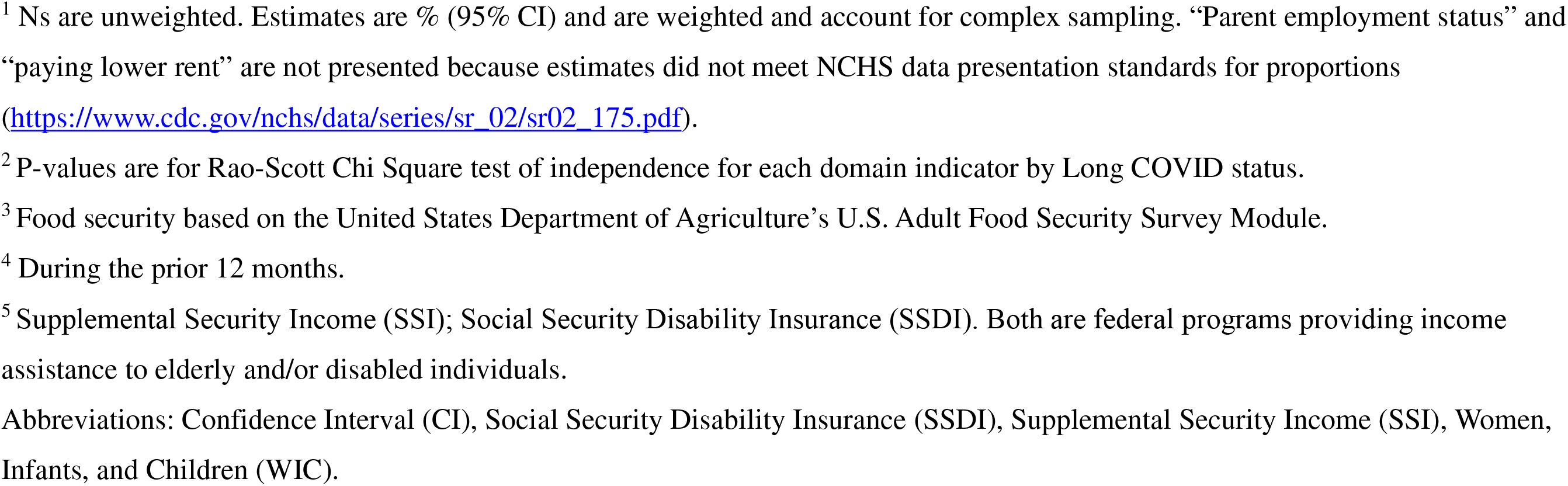
Economic Stability Domain indicators among children who ever and never had Long COVID, United States, 2022-2023. ^1^

| Characteristic | Percent of Children who ever<br>had Long COVID |  | Percent of Children who<br>never had Long COVID |  | P-value <sup>2</sup> |
| --- | --- | --- | --- | --- | --- |
|  | N | (95% CI) | N | (95% CI) |  |
| **Total** | 212 | 1.3 (1.2, 1.6) | 14,781 | 98.7 (98.4, 98.8) |  |
| **General economic stability** |  |  |  |  |  |
| Parent education |  |  |  |  | 0.0002 |
| High school diploma or less | 55 | 27.6 (21.1, 34.9) | 3,610 | 26.7 (25.5, 27.9) |  |
| Some college or associate's degree | 75 | 38.4 (30.7, 46.6) | 3,599 | 25.3 (24.4, 26.3) |  |
| Bachelor's degree or higher | 74 | 34.0 (26.8, 41.8) | 7,201 | 48.0 (46.5, 49.4) |  |
| Household food security <sup>3</sup> |  |  |  |  | 0.0006 |
| High food security | 150 | 70.0 (62.3, 77.0) | 11,932 | 80.2 (79.2, 81.1) |  |
| Marginal food security | 23 | 12.4 (7.7, 18.5) | 1,180 | 8.6 (8.1, 9.3) |  |
| Low food security | 38 | 17.6 (12.1, 24.3) | 1,439 | 11.2 (10.5, 11.9) |  |
| **Participation in social safety net programs** |  |  |  |  |  |
| Food stamps <sup>4</sup> | 55 | 27.8 (20.5, 36.0) | 2,831 | 23.0 (21.9, 24.2) | 0.18 |
| WIC benefits <sup>4</sup> | 26 | 17.2 (11.2, 24.8) | 1,501 | 13.4 (12.5, 14.3) | 0.21 |
| Free or reduced meals at school | 115 | 60.6 (52.4, 68.4) | 6,291 | 55.7 (54.1, 57.3) | 0.22 |
| Income from SSI or SSDI <sup>5</sup> | 27 | 14.0 (9.0, 20.3) | 907 | 6.4 (5.9, 6.9) | 0.0001 |
| Income from public assistance | 21 | 12.5 (7.3, 19.5) | 803 | 6.4 (5.9, 7.1) | 0.007 |
| **Housing security** |  |  |  |  |  |
| Length of time in house/apartment (years) |  |  |  |  | 0.25 |
| < 1 | 20 | 11.0 (6.3, 17.4) | 1,627 | 12.1 (11.4, 12.8) |  |
| 1-3 | 77 | 36.7 (29.5, 44.5) | 4,687 | 35.2 (34.2, 36.2) |  |
| 4-10 | 72 | 33.8 (26.8, 41.4) | 5,236 | 39.1 (38.1, 40.1) |  |
| > 10 years | 40 | 18.5 (13.1, 24.9) | 2,143 | 13.6 (13.0, 14.3) |  |
| Owned residence |  |  |  |  | 0.07 |
| Owned | 127 | 58.9 (50.7, 66.8) | 9,785 | 65.9 (64.6, 67.2) |  |
| Not owned | 84 | 41.1 (33.2, 49.3) | 4,746 | 34.1 (32.8, 35.4) |  |

<sup>1</sup> Ns are unweighted. Estimates are % (95% CI) and are weighted and account for complex sampling. “Parent employment status” and “paying lower rent” are not presented because estimates did not meet NCHS data presentation standards for proportions (https://www.cdc.gov/nchs/data/series/sr_02/sr02_175.pdf).
<sup>2</sup> P-values are for Rao-Scott Chi Square test of independence for each domain indicator by Long COVID status.
<sup>3</sup> Food security based on the United States Department of Agriculture’s U.S. Adult Food Security Survey Module.
<sup>4</sup> During the prior 12 months.
<sup>5</sup> Supplemental Security Income (SSI); Social Security Disability Insurance (SSDI). Both are federal programs providing income assistance to elderly and/or disabled individuals.
Abbreviations: Confidence Interval (CI), Social Security Disability Insurance (SSDI), Supplemental Security Income (SSI), Women, Infants, and Children (WIC).

## Discussion

Children who ever had Long COVID more frequently reported adverse SDOH than their peers who never had Long COVID. Across each of the three SDOH domains we analyzed, household-level adverse SDOH, such as low food security and financial stress, were also more common among those who ever had Long COVID compared with those who never had Long COVID. From one end, coping with adverse SDOH could increase risk of developing Long COVID or present additional barriers to managing it. From the other, coping with Long COVID could potentially exacerbate adverse SDOH, such as economic instability. Increasing awareness of the associations between SDOH and Long COVID may help healthcare providers and public health professionals identify higher-risk individuals who might benefit from additional support when they acquire covid infection and are at risk for Long COVID.

Children who ever had Long COVID more often reported ACEs than their peers who never had Long COVID. This finding is consistent with studies that have shown that ACEs are associated with poorer health outcomes.^17^ One study reported that ACEs were positively associated with increased rates of obesity, depression, and hypertension among adolescents living in urban areas.^17^ Experiencing ACEs in early childhood has also been associated with an increased risk of adverse mental, well-being, and social behavior outcomes in adolescence.^18^ A study using data from the Behavioral Risk Factor Surveillance System found that almost one in six adults reported at least four types of ACEs.^19^ The study found significant associations between experiencing ACEs and poorer health outcomes (e.g., obesity, depression), health risk behaviors (e.g., heavy drinking), and socioeconomic challenges.^19^ Moreover, such experiences have been shown to have intergenerational continuity, harming health and well-being across generations.^20^

Health insurance coverage is an important component of healthcare access and affordability. In our study, we found differences in insurance type by Long COVID status. Children who ever had Long COVID more often had public insurance than those who never had Long COVID. Public health insurance such as Medicaid and the Children’s Health Insurance Program (CHIP) provide health coverage to children at little to no cost.^21^ Although insured, these children may still face barriers to care.^22^ A study of Medicaid/CHIP programs found that 4 in 10 children with public insurance faced one or more barriers to care.^23^ Public insurance may serve as a proxy for household income or other socioeconomic factors that may affect risk of developing Long COVID.

Children who ever had Long COVID had more frequent healthcare use than their peers who never had Long COVID. Higher healthcare use among children with Long COVID may be associated with increased household medical expenditure – as healthcare-related out-of-pocket costs which have risen over the last several decades.^25^ For example, in a study of privately insured individuals, children with COVID-19 experienced higher direct costs at 1-, 3-, and 6-months post infection compared with control children.^26^In our study, nearly one in four caregivers of children with Long COVID reported problems paying medical bills compared to fewer than one in ten caregivers of children who never had Long COVID. Our findings are consistent with a study of U.S. adults that found that more than 1 in 5 adults having Long COVID had difficulty paying medical bills compared to adults not having Long COVID.^8^ Long COVID could directly contribute to financial strain through out-of-pocket payments for medical care – even among those who have insurance – or indirectly if caregivers need to take time off of work for caregiving responsibilities or other factors.^27^

Long COVID may contribute to or exacerbate existing household-level financial instability. In our study, children who ever had Long COVID resided in households that more frequently experienced income instability and financial strain than children who never had Long COVID. For example, children who ever had Long COVID more frequently resided in households receiving public assistance (e.g., SSI, SSDI) compared to children who never had Long COVID. Similarly, the prevalence of being very worried about paying future bills among caregivers of children who ever had Long COVID was double that of their peers. Relatedly, children who ever had Long COVID more often resided in single-parent households – which are typically less resourced than two-parent households and may affect a household’s ability to manage potential caregiving and financial demands of Long COVID. ^28^

### Strengths and Limitations

Strengths of this analysis include nationally representative data, including a broad range of SDOH. Some SDOH indicators, including food security, were measured using validated tools. This analysis explicitly addresses the relationship between Long COVID and SDOH in the pediatric population, which has been identified as a gap in the literature.^11^ This analysis also has several limitations. The cross-sectional design precludes us from examining causal relationships between SDOH and Long COVID. Responses are based on caregiver report, which introduces the potential for misclassification and recall bias. COVID-19 history and Long COVID may be misclassified for children with asymptomatic or mild COVID-19 illness who were not tested. Misclassification of Long COVID may be differential by child age as younger children may have more difficulty articulating their symptoms to caregivers than adolescents.^12^ This analysis examines SDOH among children who ever had Long COVID; the sample of children with current Long COVID was too small to present reportable estimates. Children who had Long COVID may or may not have had symptoms at the time of the survey. No information on symptom duration was available. Also, as previously described, not all SDOH domains are represented due to a lack of suitable indicators in the NHIS for “Neighborhood and Built Environment Domain” or the “Education Access and Quality Domain.” Because we did not adjust for multiple testing, P-values should be interpreted with caution.

### Conclusion

These findings indicate that adverse household and social conditions, such as ACEs, receiving public assistance, and low food security, were more commonly reported among children with a history of Long COVID, compared to their peers without a reported history of Long COVID. The demand of managing a chronic condition like Long COVID may potentially exacerbate poorer health and financial outcomes. Longitudinal studies are needed to clarify temporality and the role of confounding. Nevertheless, increasing awareness of the relationship between SDOH and Long COVID may help healthcare providers and public health professionals identify higher-risk children who may benefit from support related to their Long COVID care.

## Data Availability

All data are available online at

https://www.cdc.gov/nchs/nhis/documentation/

